# Seroprevalence of chikungunya virus in Colombo, Sri Lanka before the 2025 outbreak and implications for population susceptibility

**DOI:** 10.64898/2026.04.04.26350157

**Authors:** Saubhagya Danasekara, Chandima Jeewandara, Jeewantha Jayamali, Shyrar Tanussiya, Laksiri Gomes, Deneshan Peranantharajah, H.S Colambage, Maneshka Karunananda, Harshani Chathurangika, Sepali Aberathna, Thushali Ranasinghe, Madushika Dissanayake, Heshan Kuruppu, Lahiru Perera, Tibutius Thanesh, Farha Bary, Chathura Ranatunga, Dinuka Guruge, Sri Prathapan, Geetha Rathnawardana, Sanjaya Nawarathne, Enoka Liyanage, Nushara Senathilleke, Rivindu H. Wickramanayake, Navanjana Warnakulasuriya, Sahan Madusanka, Chathumini Dissanayake, Sathsara Yatiwella, Ruwan Wijayamuni, G N Malavige

**Author notes:** Corresponding author: Prof. Neelika Malavige. DPhil (Oxon), FRCP (Lond), FRCPath (UK), ^1^Institute of Allergology and Immunology, University of Sri Jayewardenepura, Gangodawila, Nugegoda, Sri Lanka and ^2^Department of Immunology and Molecular Medicine, University of Sri Jayewardenepura. Contributed equally (Saubhagya Danasekara, Chandima Jeewandara).

## Abstract

**Introduction:** Following a large chikungunya outbreak during 2006-2008, Sri Lanka did not report any outbreaks for a 16-year period until end of 2008, possibly due to population immunity. Therefore, understanding baseline immunity prior to outbreaks is crucial to inform implementation of vaccine strategies.

**Methods:** We assessed the age-stratified seroprevalence for chikungunya in an urban (n=816) and a semi-urban (n=380) community in Colombo, Sri Lanka, from September to November 2024, prior to the commencement of the large chikungunya outbreak, in December 2024. Socio-demographic, socio-economic and clinical data were collected and chikungunya specific IgG measured in serum samples.

**Results:** Of 1196 participants, 410 (34·3%) were chikungunya IgG seropositive. Seroprevalence was significantly higher in urban populations compared with semi-urban populations (39·6% vs 22·9%; p<0·001) and increased significantly with age in urban areas but not in semi-urban areas. Living in an urban area was the strongest independent risk factor of chikungunya seropositivity (aOR 7·48, 95% CI 4·05–13·81; p<0·001), consistent with the higher population density, poor housing conditions and overcrowding observed in that setting. The use of mosquito nets was independently associated with reduced risk of seropositivity (aOR 0·50, 95% CI 0·27–0·93; p=0·029). Almost no individuals aged <16 years had evidence of prior infection (0.55%), indicating minimal transmission in the preceding 16 years. In the urban cohort, seropositivity was significantly associated with diabetes, central obesity, overweight, and hypertension.

**Conclusions:** There appears to have been minimal chikungunya transmission in the 16 years preceding the 2024 outbreak, with a large population susceptible to chikungunya. Higher seroprevalence in urban populations highlights the role of population density, overcrowding, and housing conditions as key drivers of transmission.

## Introduction

Although chikungunya has caused sporadic outbreaks worldwide since its identification in Tanzania in the early 1950s, its global incidence has risen significantly in recent years ^1^. Globally the incidence increased from 0.28 per 100,000 population in 2004 to 11.13 per 100,000 population by 2024, with the most pronounced rises reported in Latin America and South-East Asia ^2^. However, there is a significant underreporting of chikungunya cases, as age-stratified seroprevalence studies and force of infection estimates, suggest that South Asia carries the highest burden ^3^. Seroprevalence studies indicate that 35 million individuals are estimated to be infected with the chikungunya virus (CHIKV) annually ^3^. Notably, there are now two vaccines that were recently approved for the prevention of chikungunya, which show seropositivity rates of >97% in vaccinees 4 weeks following vaccination ^4^ ^5^. However, due to the inconsistent nature of chikungunya outbreaks, performing clinical efficacy studies becomes difficult and therefore, vaccine efficacy rates, durability of immune responses and the proportion of population required to be vaccinated to prevent outbreaks, have been based on immunogenicity studies ^5^ ^6^. Based on estimates assuming that the vaccine provides 70% protection against disease and 40% protection against infection and an average period of protection of 5 years, it is estimated that 50% of vaccine coverage would lead to 5.8 million fewer infections an 168,000 fewer cases of chronic chikungunya illness ^3^. The vaccination coverage required to reduce the possibility of the occurrence of outbreaks, symptomatic cases and chronic illness us largely dependent on age-stratified seroprevalence data ^3^. Age-stratified seroprevalence data can inform both the proportion of the population at risk and the level of population immunity needed to prevent large-scale outbreaks, which is crucial data to inform implementation of vaccines.

Sri Lanka experienced its first large chikungunya outbreak in years 2006 to 2008, reporting over 37,000 suspected cases ^7^. Although Sri Lanka did not report any chikungunya cases until the next large outbreak in the late 2024, there is evidence that sporadic chikungunya cases were present ^8^. The year 2006 to 2008 outbreak in Sri Lanka was likely to linked to a series of large outbreaks that occurred in the Indian Ocean islands that preceded this period ^7^. Studies of febrile patients presenting to clinics in Sri Lanka immediately prior to the first outbreak showed no evidence of current or past CHIKV infection ^9^. Therefore, lack of population immunity to the CHIKV may have led to Sri Lanka experiencing the large outbreak in 2006 to 2008. The absence of outbreaks in Sri Lanka for 16 years, until late 2024, may be attributable to population immunity limiting transmission. Therefore, defining the level of population immunity required to prevent outbreaks is essential to inform the implementation of chikungunya vaccination strategies. Furthermore, given evidence that the CHIKV has undergone mutations at key neutralizing antibody binding sites ^10^, it is important to assess whether the emergence of such variants could lead to outbreaks even in the presence of substantial population immunity.

Sri Lanka experienced a large chikungunya outbreak that emerged in the latter part of 2024, with cases mainly reported in adults between 40 to 60 years of age ^11^ ^12^. Females between ages 40-60 were predominantly affected with a significantly higher symptom scores and disability at presentation and a significantly delayed resolution of symptoms compared to other age groups ^12^. As the last chikungunya outbreak was reported 16 years ago, the most susceptible group is likely to have been children <16 years of age, with individuals >16 years of age being equally susceptible to infection. Although arthritis is more prominent in adults with chikungunya than children ^13^, the vast majority of infections in children have shown to be symptomatic ^14^. Therefore, although there could be multiple reasons for increased symptomatic illness predominantly in the 40-60 year old age group, it would be important to understand how population immunity affects transmission of chikungunya. In this study, we determined the age-stratified seroprevalence rates for chikungunya in a urban and a semi-urban population in Colombo, Sri Lanka before the occurrence of the large outbreak towards December 2024. Therefore, establishing the threshold of population immunity required to prevent outbreaks is critical for guiding the deployment of chikungunya vaccines.

## Methods

We recruited 1196 individuals aged 5 to 86 years (mean 44.2, SD ± 20.9) from a large cohort from individuals from an urban and a semi-urban community in Colombo, Sri Lanka during the months of September to early November 2024. The urban study area (Kirulapone) covers 1.23 km² (123 hectares) with a population of 17,026, with a population density of 13,842 persons/km² based on the 2024 population census ^15^ (Figure 1). The semi-urban study area comprised two Grama Niladhari administrative divisions (smallest administrative unit), namely the Rattanapitiya and Egodawatta areas within the Boralesgamuwa Urban Council. This area covers a combined area of 1.5 km² with a population of 8,913 and a population density of 5942 persons/km² according to the 2024 population census ^15^ (Figure 1).

**Figure 1.**
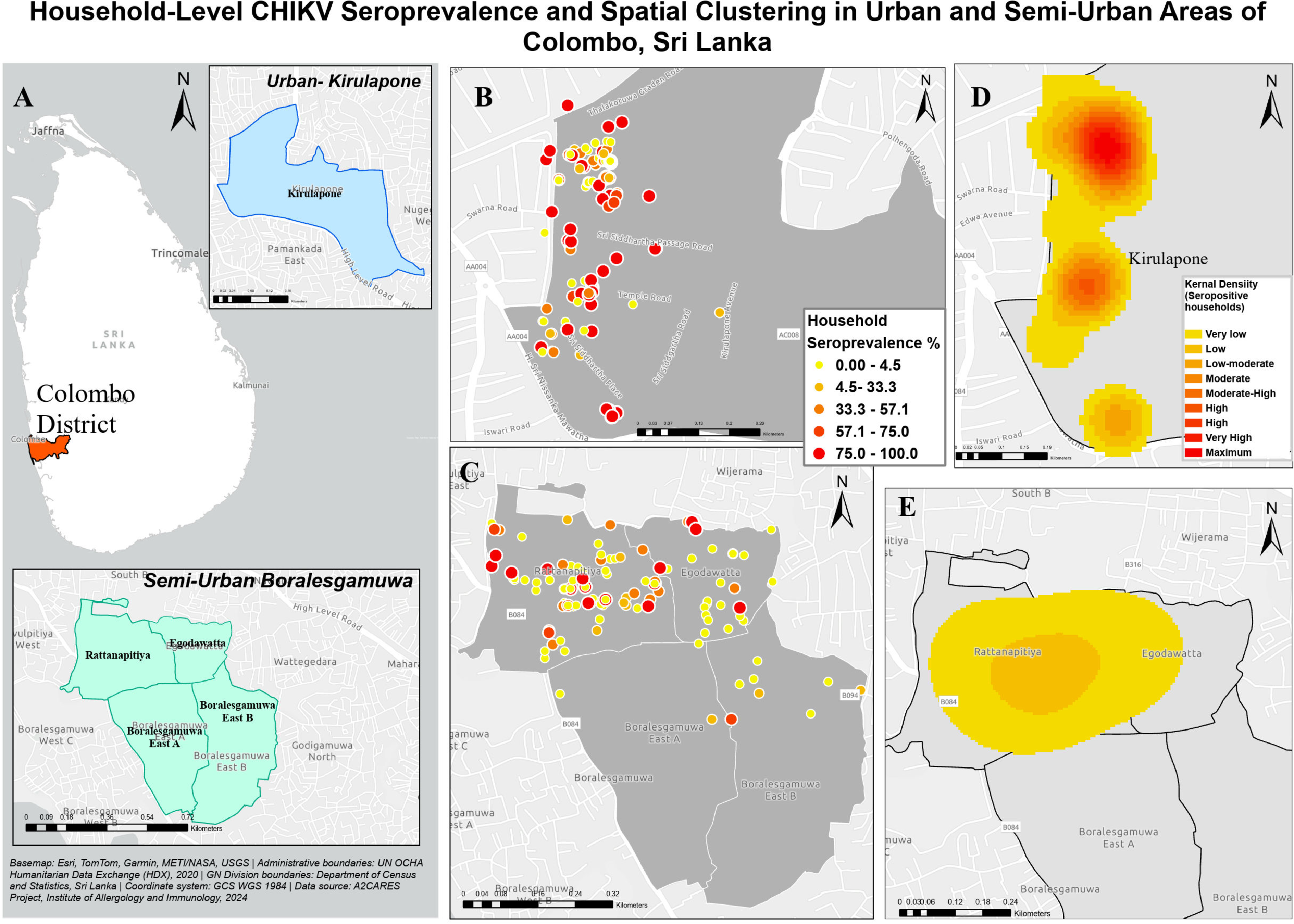
Household-level seroprevalence chikungunya and spatial clustering in Colombo, Sri Lanka. (A) The Study site locations within Sri Lanka with the upper inset showing the Kirulapone urban study area (Colombo Municipal Council with a population density 13,842 persons/km²) and the lower inset showing the Rattanapitiya and Egodawatta semi-urban study area (Boralesgamuwa Urban Council with a population density of 5942 persons/km²). The Colombo District is highlighted in red. (B–C) indicate household CHIKV seroprevalence (%) in urban Kirulapone (B) and semi-urban Boralesgamuwa (C) areas. The symbol colour indicates household seroprevalence and the symbol size is proportional to household members tested. (D–E) Kernel density estimation of seropositive households in Kirulapone (D) and Boralesgamuwa (E). The colour intensity represents spatial clustering of seropositivity weighted by seropositive individuals per household. Basemap: Esri, TomTom, Garmin, METI/NASA, USGS. Boundaries: UN OCHA HDX 2020; Department of Census and Statistics, Sri Lanka. Data: A2CARES Project 2024. CHIKV=chikungunya virus.

Individuals living in the randomly selected households were invited to participate in the study and venous blood samples were collected following informed written consent. Serum was separated and stored at -80 C. Demographic data and socio-economic data were collected using an interviewer administered questionnaire from the participants to assess any relationship between, housing conditions and socio-economic status with CHIKV-IgG seropositivity. Only 412/816 from the urban area and only 188/380 from the semi-urban area, responded to questions regarding the level of education, while only 491/816 and 280/380 from the semi-urban area responded to questions regarding occupation.

As individuals with comorbidities have shown to have a higher incidence of severe chikungunya illness ^16^ ^17^, we also assessed the presence of diabetes and central obesity in the cohort. Fasting blood samples were obtained from participants who were >18 years of age and they were considered to have diabetes, if they were already on treatment for diabetes or if they had a FBS of ≥100 mg/dL ^18^. Individuals were considered to have hypertension if they were already diagnosed with hypertension or if a systolic blood pressure of ≥130 and/or diastolic blood pressure of ≥85 mm Hg was detected measured 1 hour apart ^18^. The waist circumference was measured using a non-stretchable tape at the midpoint between the lowest rib and the iliac crest and the hip circumference was measured at the level of the widest lateral extension of the pelvis. Individuals >18 years of age, with a waist circumference over the population cut-off values (≥80cm for females and ≥90cm for males), were considered to have central obesity ^18^. Body mass index (BMI) was calculated as weight divided by height squared (kg/m²) and individuals with a BMI of >23.9 was classified as overweight based on the South Asian–specific cut-off values ^19^.

### Ethics approval

Ethics approval was obtained from the Ethics Review Committee of the University of Sri Jayewardenepura. Informed written consent was obtained from all study participants. Informed written consent was obtained from the parents or guardians in children <18 years of age and assent was obtained from children aged 12-18 years. Administrative clearance was obtained from the Ministry of Health Sri Lanka. The study was carried out conforming to the principles embodied in the Declaration of Helsinki.

### Detection of Chikungunya virus (CHIKV) specific IgG antibodies in the population

Serum samples were tested for CHIKV-specific IgG antibodies using the SERION ELISA classic Chikungunya Virus Igg kit (Institut Virion\Serion GmbH, Germany). The assay was carried out and the results interpreted according to the manufacturer’s instructions.

### Statistical analysis

All analyses were done using Python (version 3.13; pandas, NumPy, SciPy, and Matplotlib, statsmodels). Continuous variables are presented as mean (SD) or median (IQR), and categorical variables as counts (percentages). Differences between Urban and Semi-Urban populations were assessed using the Mann–Whitney U test for continuous variables and the Pearson chi square (χ²) test (with Yates’ continuity correction for 2×2 tables) for categorical variables, with 95% CIs reported where appropriate. Among participants aged 18 years or older, associations between seropositivity and metabolic risk factors, including diabetes mellitus and central obesity, were evaluated using χ² tests. The association between age and seroprevalence was assessed separately for Urban and Semi-Urban populations using Spearman’s rank correlation coefficient. To identify independent risk factors for CHIKV seropositivity, a multivariable logistic regression model was fitted among participants aged ≥18 years with complete data. The outcome variable was CHIKV-specific IgG seropositivity (seropositive vs seronegative; borderline results excluded). Candidate variables were first screened using univariable logistic regression; those with p<0·20 were considered for inclusion in the multivariable model alongside variables retained on the basis of biological plausibility. Model fit was assessed using the McFadden pseudo R² and Akaike information criterion (AIC) using statsmodels package (version 0.14.4).

Spatial analyses and maps were produced using ArcGIS Pro (version 3.6) with kernel density estimation used to visualize spatial distribution patterns. Household GPS coordinates were recorded during field data collection. Spatial distribution of CHIKV household seroprevalence was analyzed at the household level. Each household was assigned a seroprevalence value calculated as the proportion of seropositive individuals among all tested household members. Kernel density estimation (KDE) was performed to identify spatial clustering of seropositive households using the Kernel Density tool in ArcGIS Pro (Version 3.6, Esri Inc., Redlands, CA, USA) with the Spatial Analyst extension. KDE was weighted by the number of seropositive individuals per household (Population field = Positives). A search radius of 0.001 decimal degrees (approximately 100 metres) was applied for the urban study area and 0.003 decimal degrees (approximately 300 metres) for the semi-urban study area, reflecting the differing spatial scales of household clustering in each setting. Output cell size was set to 0.0001 decimal degrees. All spatial analyses were conducted using the Geographic Coordinate System WGS 1984.

### Patient and Public involvement in research

Patients and the public were not involved in the study design or in the analysis of results. Community leaders were initially informed of this longitudinal study titled ‘Asian American Centres for Arbovirus Research and Enhanced Surveillance (A2CARES) community study, which was initiated to study arbovirus surveillance and immune responses in dengue and other arboviruses, prior to participant recruitment. Several workshops were held in the respective communities to educate the public residing in areas regarding the study objectives, with the study team answering all questions. Participant information sheets were distributed at least one week prior to recruitment to the study, so that participants had time to consider if they would join and to ask relevant questions.

## Results

The demographic characteristics and socio-economic characteristics of individuals from the urban are semi-urban areas are shown in Table 1. The semi-urban cohort was significantly older, significantly more likely to have received education of college level and beyond with differences in the type of employment undertaken. The number of children per household was significantly higher in the urban area, with significantly small house area and number of rooms used for sleeping. Further, the population density was significantly higher in the urban area, and housing conditions, material used for housing was significantly different. The average area of an urban house was 3.23 (± 4.30) perches with an average of 4.41 (± 2.23) individuals living in each household. The average area of a house was 11.74 (± 9.33) perches with an average of 4.19 (± 1.94) individuals living in each household. 816 (68.2%) individuals were recruited from the urban areas whereas 380 (31.8%) were recruited from the semi-urban areas. The population density in the urban area (13,842 persons/km²) was substantially higher than the semi-urban areas (5942 persons/km²) representing a 2.33-fold difference indicating a large contextual difference between the two study sites.

**Table 1:** Socio-demographic and socio-economic characteristics of the urban and semi-urban population. *p-value^†^ from Chi-square test; p-value **^‡^** from Man-Whitney U test at 5% significance level*.

| Characteristic | Urban Household<br>Population N= 816<br>Household n= 311 | Semi-urban<br>Household<br>Population N=380 | P value |
| --- | --- | --- | --- |
|  |  | **Household n=266** |  |
| Age | 41.11 ± 20.95 | 50.94 (19.24) |  |
| Mean ± SD [Median, IQR] | [43.5, 19-58] | [54,40-67] | <0.001<sup>‡</sup> |
| Females n (%) | 483 (59.2) | 230 (60.5) | 0.71<sup>†</sup> |
| Males n (%) | 333 (40.8) | 150 (39.5) |  |
| **Education Level** |  |  |  |
| Primary | 60 (14.6%) | 6 (3.2%) | <0.001<sup>†</sup> |
| Junior secondary | 83 (20.2%) | 6 (3.2%) |  |
| Senior secondary | 195 (47.3%) | 60 (31.9%) |  |
| Collegiate | 58 (14.1%) | 87 (46.3%) |  |
| Tertiary | 16 (3.8%) | 29 (15.4%) |  |
| **Occupation** |  |  |  |
| Formal employment | 72 (14.7%) | 53 (18.9%) | <0.001<sup>†</sup> |
| Informal employment | 124 (25.3%) | 39 (13.9%) |  |
| Unemployed | 171 (34.8%) | 99 (35.4%) |  |
| Student | 99 (20.2%) | 39 (13.9%) |  |
| Retired | 25 (5.1%) | 50 (17.9%) |  |
| **Household Composition** |  |  |  |
| Adults per household | 3.24 ± 1.39 | 3.25 ± 1.58 | 0.983<sup>‡</sup> |
|  | [3.0, 2.0–4.0] | [3.0, 2.0–4.0] |  |
| Children per household | 1.18 ± 1.40 | 0.94 ± 1.10 | 0.022<sup>‡</sup> |
|  | [1.0, 0.0–2.0] | [1.0, 0.0–2.0] |  |
| Total household members<br>(Population) | 4.41 ± 2.23<br>[4.0, 3.0–5.0] | 4.19 ± 1.94<br>[4.0, 3.0–5.0] | 0.203<sup>‡</sup> |
| Sleeping rooms | 2.29 ± 2.68<br>[2.0, 2.0–2.0] | 2.91 ± 1.39<br>[3.0, 2.0–4.0] | < 0.001<sup>‡</sup> |
| House area (perches) | 3.23 ± 4.30<br>[2.5, 2.0–2.5] | 11.74 ± 9.33<br>[10.0, 6.0–14.0] | < 0.001<sup>‡</sup> |
| Housing Material |  |  |  |
| Floor material, n (%) |  |  |  |
| Ceramic | 136 (43.7%) | 172 (64.7%) | < 0.001<sup>†</sup> |
| Concrete | 158 (50.8%) | 83 (31.2%) |  |
| Carpet | 17 (5.5%) | 9 (3.4%) |  |
| Mud brick | 0 (0.0%) | 2 (0.8%) |  |
| Wall material, n (%) |  |  |  |
| Concrete | 310 (99.7%) | 265 (99.6%) | 0.363<sup>†</sup> |
| Wood | 1 (0.3%) | 0 (0.0%) |  |
| Other | 0 (0.0%) | 1 (0.4%) |  |
| Roof material, n (%) |  |  |  |
| Metal (Zinc) | 221 (71.1%) | 154 (57.9%) | < 0.001<sup>†</sup> |
| Concrete | 82 (26.4%) | 39 (14.7%) |  |
| Wood | 4 (1.3%) | 24 (9.0%) |  |
| Clay | 0 (0.0%) | 8 (3.0%) |  |
| Garbage collection frequency, n (%) |  |  | <0.001<sup>†</sup> |
| Daily | 4 (1.3%) | 5 (1.9%) |  |
| 2 or 3 times per week | 252 (81.0%) | 74 (27.8%) |  |
| Once a week | 53 (17.0%) | 171 (64.3%) |  |
| Once every 2 weeks | 2 (0.6%) | 8 (3.0%) |  |
| Once every month | 0 (0.0%) | 3 (1.1%) |  |
| No garbage collection service | 0 (0.0%) | 3 (1.1%) |  |

### Age stratified seroprevalence rates of CHIKV seropositivity rates in urban and semi-urban areas

Of the 1196 individuals, 410 (34.3%) tested positive for CHIKV-specific IgG with 6 individuals showed borderline results. CHIKV-specific IgG antibodies were not detected in any of the children <16 years of age from the semi-urban area, whereas 2 (0.55%) from the urban area tested positive for CHIKV-specific IgG (Table 2). After 16 years of age, the age stratified seroprevalence rates, significantly increased in the urban area (Spearman’s r=0.93, p=0.003), but not in the semi-urban area (Spearman’s r=0.61, p=0.148) (Figure 2 and Table 2). There were no differences in seropositivity rates in females vs males. Interestingly, the age stratified seropositivity rates were significantly higher in all age groups older than 25 years of age in the urban area compared the semi-urban area.

**Figure 2.**
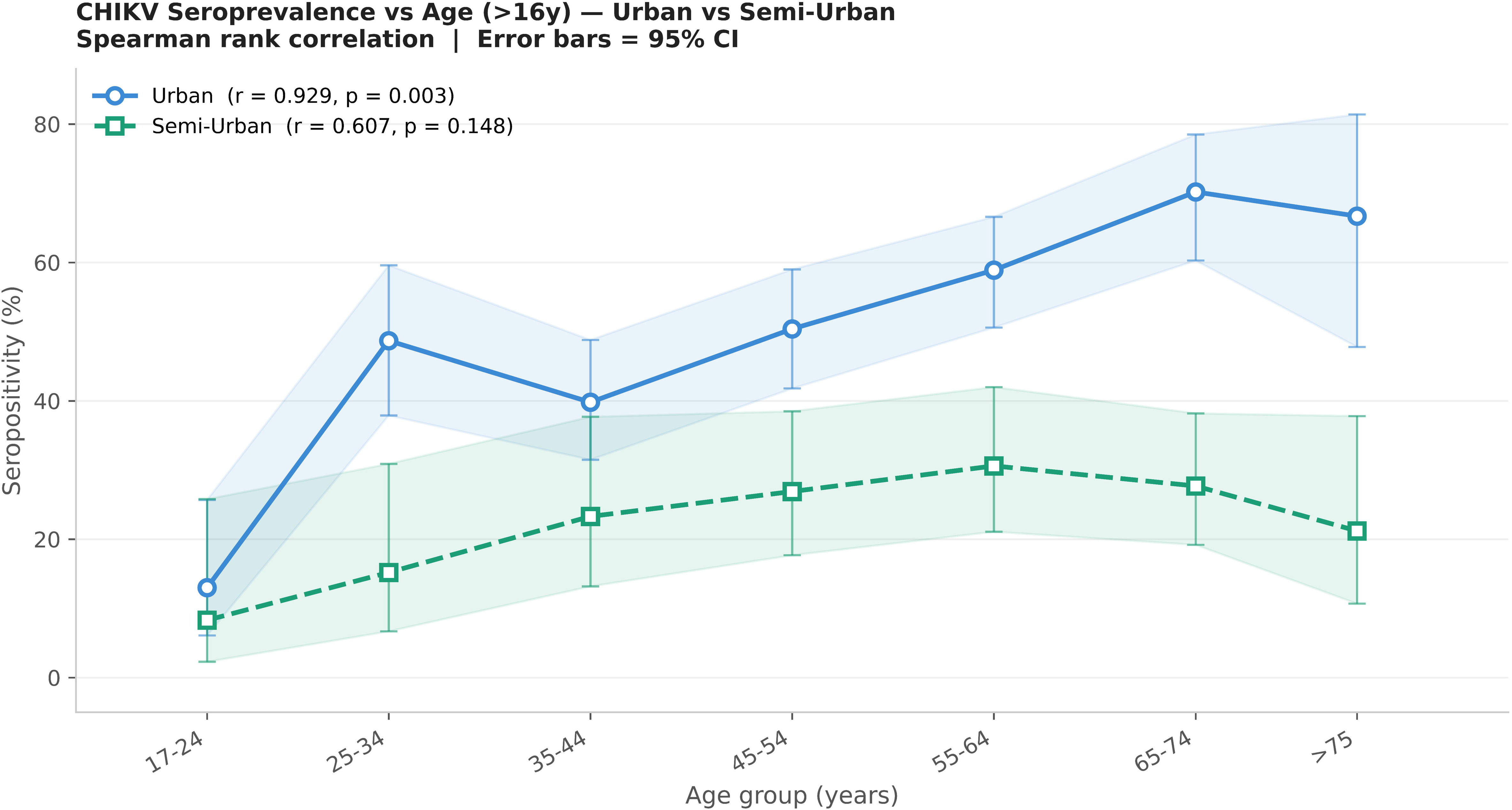
CHIKV-specific IgG seroprevalence by age group (>16 years) in urban and semi-urban study populations in Sri Lanka. Seroprevalence (%) is plotted for adults aged >16 years across seven age groups in urban (Kirulapone) and semi-urban (Boralesgamuwa). Error bars represent 95% confidence intervals. Spearman rank correlation between age group midpoint and seroprevalence was significant in the urban population (r = 0.929, p = 0.003) but not in the semi-urban population (r = 0.607, p = 0.148), indicating a significant positive monotonic increase in CHIKV seropositivity with age in the urban area only.

**Figure 3.**
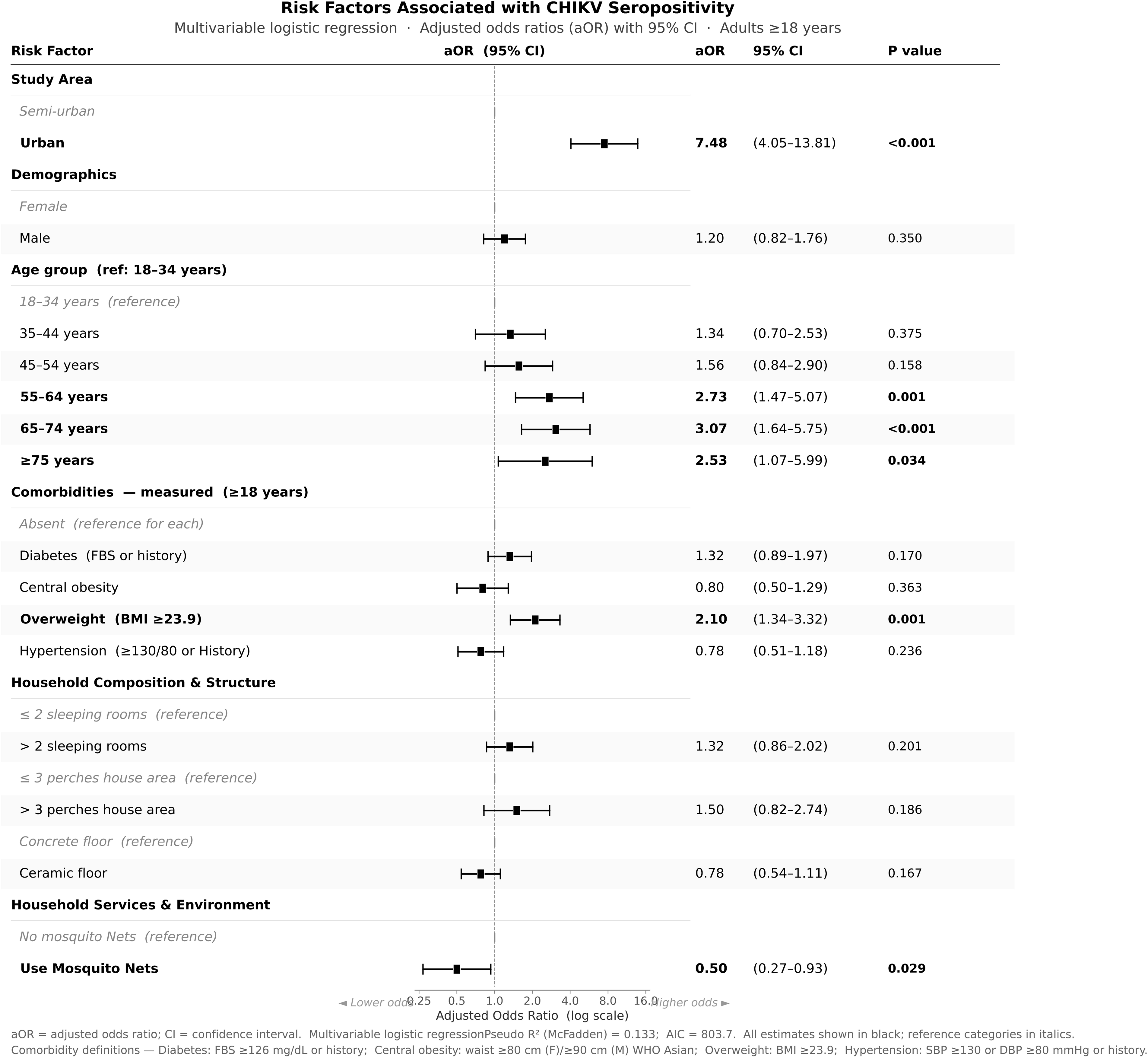
Adjusted odds ratios for risk factors associated with CHIKV-specific IgG seropositivity. Forest plot showing adjusted odds ratios (aOR) with 95% confidence intervals (CI) from multivariable logistic regression among adults aged ≥18 years (n=652 complete cases; 279 seropositive, 373 seronegative). The model was adjusted simultaneously for study area, gender, age group, diabetes, central obesity, overweight, hypertension, sleeping rooms, house area, floor material and mosquito screens. Age groups are referenced against the 18–34 years group. The vertical dashed line represents an aOR of 1·0 (no association). Bold labels and values indicate associations reaching statistical significance (p<0·05). aOR=adjusted odds ratio. CI=confidence interval. BMI=body mass index.

**Table 2:**
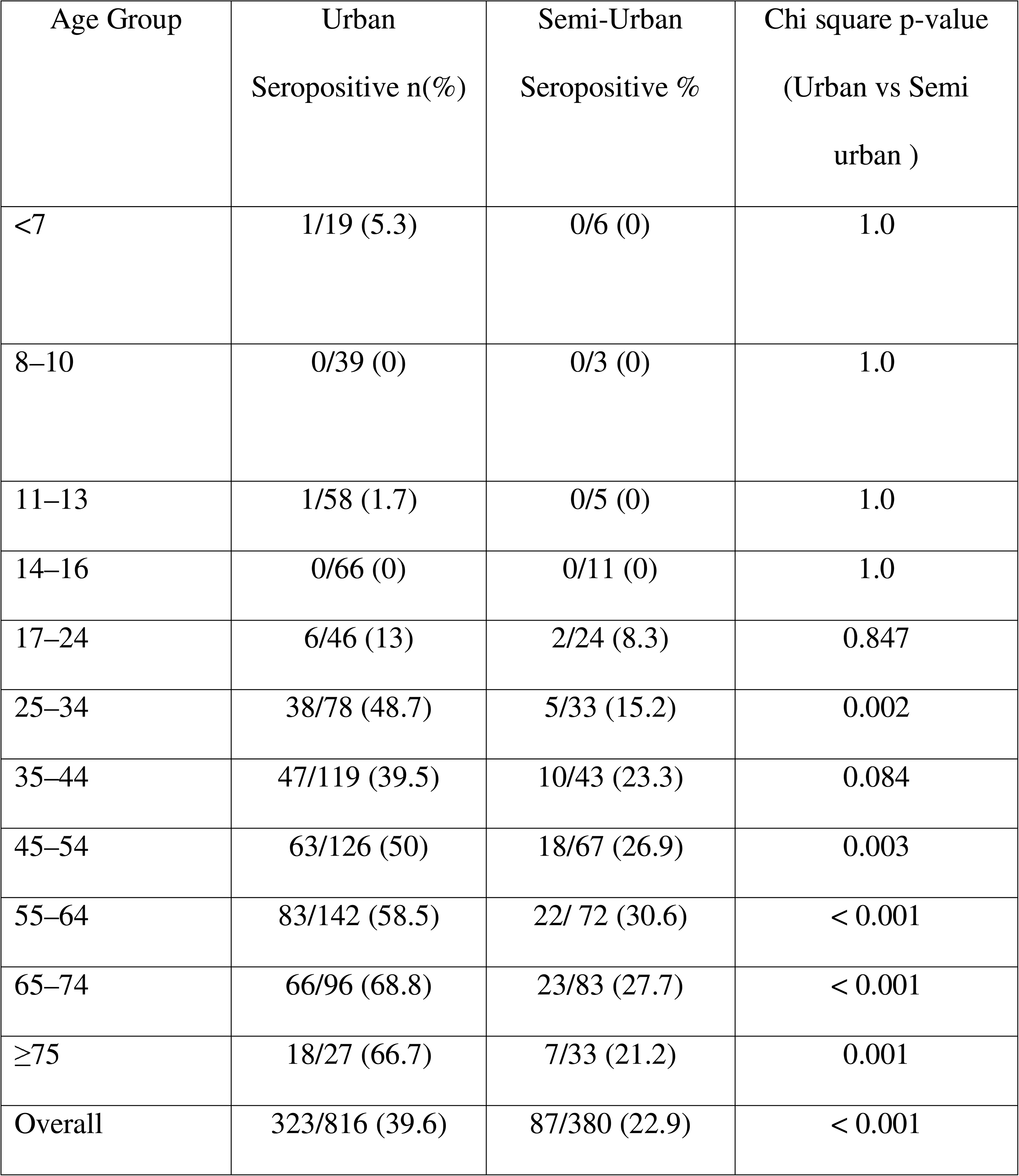
Age stratified seropositivity rates of CHIKV-specific IgG antibodies in different age groups in an urban and a semi-urban population in Sri Lanka.

| Age Group | Urban<br>Seropositive n(%) | Semi-Urban<br>Seropositive % | Chi square p-value<br>(Urban vs Semi<br>urban ) |
| --- | --- | --- | --- |
| <7 | 1/19 (5.3) | 0/6 (0) | 1.0 |
| 8–10 | 0/39 (0) | 0/3 (0) | 1.0 |
| 11–13 | 1/58 (1.7) | 0/5 (0) | 1.0 |
| 14–16 | 0/66 (0) | 0/11 (0) | 1.0 |
| 17–24 | 6/46 (13) | 2/24 (8.3) | 0.847 |
| 25–34 | 38/78 (48.7) | 5/33 (15.2) | 0.002 |
| 35–44 | 47/119 (39.5) | 10/43 (23.3) | 0.084 |
| 45–54 | 63/126 (50) | 18/67 (26.9) | 0.003 |
| 55–64 | 83/142 (58.5) | 22/ 72 (30.6) | < 0.001 |
| 65–74 | 66/96 (68.8) | 23/83 (27.7) | < 0.001 |
| ≥75 | 18/27 (66.7) | 7/33 (21.2) | 0.001 |
| Overall | 323/816 (39.6) | 87/380 (22.9) | < 0.001 |

### Association of comorbidities with CHIKV-IgG seropositivity in the population

As individuals with diabetes and other comorbidities are more likely to develop severe illness during acute chikungunya and also at a higher risk of developing chronic arthritis ^16^ ^17^, we assessed CHIKV-IgG seropositivity rates in those with diabetes and central obesity in individuals ≥18 years of age in both the urban and the semi-urban cohort. The prevalence of comorbidities and association with CHIKV seropositivity rates are shown in Table 3. In the urban area, individuals with diabetes, central obesity, those who were overweight or had hypertension were significantly more likely to be seropositive for the CHIKV. Although a similar trend was observed for CHIV seropositivity and comorbidities, these differences were not significant in the semi-urban area.

**Table 3:** Association of comorbidities with CHIKV seropositivity rates in the urban and the semi-urban population ≥ 18 years.

| Characteristic | Urban<br>CHIKV-seropositive<br>n=320 | Urban<br>CHIKV-seronegative<br>n=298 | Chi<br>square P<br>value | Semi-urban<br>CHIKV-seropositive<br>n=86 | Semi-urban<br>CHIKV-seronegative<br>n=264 | Chi<br>square P<br>value |
| --- | --- | --- | --- | --- | --- | --- |
| Diabetes | 88 (27.5%) | 53 (17.8%) | 0.004 | 25 (29.1%) | 57 (21.6%) | 0.155 |
| Central<br>obesity | 188<br>(58.8%) | 146 (48.9%) | 0.037 | 43 (50%) | 130 (49.2%) | 0.465 |
| Overweight<br>BMI ≥ 23.9 | 207<br>(64.7%) | 163 (55.5%) | 0.019 | 55 (63.9%) | 139 (52.6%) | 0.060 |
| Hypertension | 234<br>(73.1%) | 192 (64.4%) | 0.020 | 54 (62.8%) | 163 (61.7%) | 0.862 |

### Multivariable analysis of risk factors for CHIKV-seropositivity

As urbanicity, housing conditions, overcrowding, age, and comorbidities appear to affect CHIKV-IgG seropositivity, we then carried out a multivariable analysis to determine risk factors associated with CHIKV-IgG seropositivity. The model included 652 adults aged ≥18 years with complete data (279 seropositive, 373 seronegative individuals) and the model was adjusted simultaneously for study area, gender, age group, comorbidities, and household-level variables (McFadden pseudo R²=0·133; AIC=803·7). Living in an urban area was the strongest independent predictor of CHIKV seropositivity (aOR 7·48, 95% CI 4·05–13·81; p<0·001), consistent with the higher population density and overcrowding observed in that setting. Compared with those aged 18–34 years, individuals aged 55–64 years (aOR 2·73, 95% CI 1·47–5·07; p=0·001), 65–74 years (aOR 3·07, 95% CI 1·64–5·75; p<0·001), and ≥75 years (aOR 2·53, 95% CI 1·07–5·99; p=0·034) had significantly higher risk of seropositivity. Among comorbidities, being overweight (BMI ≥23·9 kg/m²) was independently associated with increased risk of seropositivity (aOR 2·10, 95% CI 1·34–3·32; p=0·001), whereas the associations with diabetes, central obesity, and hypertension were not significant. The use of mosquito nets was independently associated with reduced risk of seropositivity (aOR 0·50, 95% CI 0·27–0·93; p=0·029).

## Discussion

In this study we determined the age stratified seroprevalence of chikungunya in an urban and semi-urban area in Colombo, Sri Lanka prior to the large chikungunya outbreak that commenced in December 2024. We found that none of the children <16 years were CHIKV-IgG seropositive in the semi-urban area, 2 (0.55%) from the urban area were seropositive. The previous large outbreak of chikungunya was seen in Sri Lanka during 2006 to 2008 (16 years ago) ^7^ ^9^. The lack of evidence of prior CHIKV infection in children under 16 years of age over this 16-year period indicates minimal transmission in Sri Lanka, despite occurrence of several outbreaks observed in several Indian states during this time period ^20^ ^21^. After the chikungunya outbreaks that were seen from 2005 to 2008 in South Asia, India, Bangladesh and Pakistan all experienced large outbreaks during 2016 to 2017 ^20^ ^22^ ^23^. The reasons for absence of outbreaks in Sri Lanka is not clear. However, our seroprevalence data indicate that silent transmission was unlikely.

Our data shows that CHIKV seropositivity rates markedly increased after 17 years of age. The CHIKV-IgG seropositivity rates were significantly higher in all age groups in the urban area, compared to semi-urban areas, implying higher transmission rates in urban areas. Furthermore, while there was a significant increase in the age stratified CHIKV-seropositivity in urban areas, no such increase was seen in the semi-urban area. The urban area had a significantly higher population density, smaller household sizes with more individuals sharing sleeping spaces. Furthermore, education level, occupations and housing characteristics were significantly different between the urban and semi-urban areas, with urban areas more likely to be comprised of marginalized communities. Our multivariate analysis also showed that living in an urban area was the most significant risk factor for CHIKV-seropositivity, highlighting the importance of factors such as overcrowding, population density and housing characteristics as potential drivers of transmission. Interestingly, use of mosquito nets was independently associated with reduced risk of seropositivity, although the Aedes species of mosquitoes are considered as ‘day biters’, and mosquito nets not considered to be effective. Therefore, in addition to climatic factors, rapid unplanned urbanization and overcrowding seen in the last two decades in many areas in Asia and Latin America, is likely to have significantly contributed to the marked increase of many mosquito borne infections such as dengue, chikungunya and zika globally ^24^. Addressing the growing threat of arboviral diseases will require sustainable strategies to reduce overcrowding and improve housing conditions in many cities across Asia and Latin America.

Sri Lanka experienced a large chikungunya outbreak that commenced at the end of 2024, with no data of the possible number of infections that occurred since December 2024. However, it is possible that Sri Lanka reported the highest incidence of chikungunya in the region in 2025, as 75/112 (66.9%) travel associated chikungunya infections reported by British tourists in 2025, were following travel to Sri Lanka ^25^. 60% of the urban population in Colombo and 77.1% of the semi-urban population in Colombo did not have evidence of CHIKV-specific IgG at the end of 2024, with possibly lower seropositivity rates in other regions in Sri Lanka. Although Sri Lanka did not experience any chikungunya outbreaks during 2008 to 2024 (16 years), the seropositivity rates reported in our study among adults, in higher than many regions in India and Bangladesh ^26^ ^27^. The CHIKV seropositivity rates were >50% in those >45 years of age in the urban area. Our multi-variant analysis showed that those >50 years of age in 2024, were significantly more likely to be seropositive for CHIKV. Although high seropositivity was reported in those >50 years of age, the highest number of cases in the 2025 chikungunya outbreak was also reported in these age groups ^11^ ^12^. Therefore, the reasons for higher number of reported cases in this age group, despite relatively higher seropositivity rates should be further investigated. In order to implement chikungunya vaccines to prevent transmission, it would be important to determine the seropositivity rates in a population and in different age groups that prevent transmission of infection.

As individuals with comorbidities are more likely to develop severe chikungunya illness and complications ^16^ ^17^, we also determined seropositivity rates in those with diabetes, obesity and hypertension. We found that in the urban area, overweight individuals, or individuals with diabetes, central obesity or hypertension were significantly more likely to be seropositive for CHIKV. Although these differences did not reach statistical significance in the semi-urban area, this could be due to lower overall seropositivity rates. Although disease severity is increased in individuals with comorbidities, this does not account for higher seropositivity rates observed and there is no clear explanation for this observation.

In summary, this is the first study of age-stratified seroprevalence in Colombo, Sri Lanka comparing age stratified seroprevalence in an urban (predominantly marginalized community) and a semi-urban community. Prior to the large outbreak that commenced in December 2024, Sri Lanka had not experienced a chikungunya outbreak for 16 years. In keeping with these epidemiological observations only 0.55% of those <16 years were seropositive to CHIKV. In those >17 years of age, the age-stratified seroprevalence rose significantly in the urban but not semi-urban area, and the CHIKV seropositivity rates were significantly higher in the urban population in all ages, suggesting higher transmission rates, also highlighting the importance of overcrowding, population density and housing characteristics in CHIKV transmission.

## Data Availability

All data produced in the present work are contained in the manuscript

## Acknowledgments

We are grateful for all the public health midwives and public health inspectors of both the urban study area (Kirulapone) and the semi-urban study area the Rattanapitiya and Egodawatta areas within the Boralesgamuwa Urban Council for their assistance in the study in conducting public education and engagement activities.

